# Trends and factors associated with initiation of HIV treatment and uptake of viral load testing among PLHIV in Jamaica

**DOI:** 10.1101/2022.03.08.22271856

**Authors:** Anya Cushnie, Ralf Reintjes, Miia Artama, J. Peter Figueroa

## Abstract

**Introduction:** Jamaica did not achieve the UNAIDS 90-90-90 targets in 2020. This study aims to examine trends and factors associated with uptake of HIV treatment and viral load testing among people living with HIV (PLHIV) in Jamaica, to make recommendations for improving patient management and outcomes.

**Methods:** This secondary analysis uses patient-level data from the National Treatment Service Information System. The baseline sample is PLHIV initiating anti-retroviral treatment (ART) between January 2015-December 2019. Descriptive statistics are used to summarize demographic and clinical variables. Multivariable logistic regression is used to assess factors associated with ART initiation (31+ days vs. same day) and viral load testing uptake (viral load test vs. no test), using categorical variables for age group, gender and regional health authority. Adjusted odds ratios and 95% confidence intervals are reported.

**Results:** Same day ART initiation increased by 72% over 5 years. The coverage of 1^st^ viral load test was 90% but declined to 79% for the 2^nd^ test. Testing occurred mostly after 0-6 months on ART (n=3047, 55%) and uptake was highest in the South-East Region (n=2885, 53%). Those virally suppressed were significantly more likely to have same day ART initiation compared to those non-suppressed (aOR=1.58, CI=1.43-1.75). Males were significantly more likely to have same day ART initiation (aOR=1.46, CI=1.32-1.62) but no first viral load test (aOR=1.55, CI=1.27-1.90) compared to females.

**Conclusion:** The goal of immediate ART initiation is increasingly being met and is significantly associated with viral suppression at the first viral load test. Males were less likely to have a viral load test after ART initiation. A qualitative assessment should be conducted to understand important challenges faced to access routine viral load testing, followed by implementation of differentiated service care models, targeting males.

## Introduction

At the end of 2019, Jamaica had an adult HIV prevalence of 1.5% and an estimated number of 32,000 persons living with HIV (PLHIV)[1]. With regards to the second and third UNAIDS 90-90-90 targets which aim to retain 90% of PLHIV diagnosed on antiretroviral therapy (ART) and ensure that 90% of people receiving ART are virologically suppressed by 2020[2], the country has achieved 44% ART retention and 35% virological suppression[3]. Jamaica is lagging considerably and did not achieve the UNAIDS targets in 2020[4].

The Government of Jamaica has provided public access to HIV treatment since 2004[1]. Jamaica also implemented the WHO Treat All strategy in January 2017[5] and a key objective is immediate initiation of ART following HIV diagnosis, regardless of clinical status[6,7]. Despite these efforts to broaden access, a vast majority of PLHIV are still not on treatment[8]. A number of randomized trials have demonstrated multiple benefits of rapid ART initiation compared to later ART start, including improved ART uptake, increased retention in care, higher rates of viral suppression and reduced risk of mortality[9,10].

There are several barriers to scale up of viral load (vl) testing in low and middle income countries, such as limited laboratory resources, missed appointments or infrequent measurements[11]. Viral load testing in Jamaica has reportedly been suboptimal due to non-adherence to testing protocols as well as laboratory-based challenges[1]. Viral load testing coverage declined from 53% in December 2019 to 45% in June 2020 [12], three months after COVID-19 protocols, including stay at home orders, were implemented locally. The viral load in patients receiving ART is the best predictor of treatment outcomes[13] but assessing patients with missing test results is also useful to identify appropriate factors that may impede uptake of viral load testing and achieving the suppression target as well as improve the quality of patient management.

The Ministry of Health and Wellness (MoHW) has earmarked the year 2025 for achievement of the 90-90-90 targets[1]. For this to occur, an analysis of factors associated with ART and viral load uptake was recommended in the recent National HIV Strategic Plan. This study aims to describe PLHIV who accessed ART and viral load testing and assess factors associated with uptake of ART and viral load testing.

## Methods

This secondary analysis uses the National Treatment Service Information System (TSIS2). TSIS2 is a centralized case-based database, governed by the MoHW and currently being used by all HIV treatment sites in Jamaica. TSIS2 stores patient level demographic and clinical data using over 100 variables.

The MoHW Internal Review Board provided ethical approval (Study No: 2017/20). The dataset was extracted by the MoHW and fully anonymized before sharing with the study investigator, as a result patient consent was waived.

### Sample Population

The baseline sample was a total of 8363 PLHIV who initiated HIV treatment between January 2015 and December 2019. For the analytical sample (n=8149), persons in the age group <15 years (n= 215) were excluded because of the small sample size compared to other age categories. To assess viral load testing uptake, the sample included only those eligible within the study period.

### Data Analysis

Descriptive statistics were used to summarise demographic and clinical variables. The categorical factors evaluated included age group, gender, location by regional health authority (RHA) and first viral load test (vl) status (suppressed or non-suppressed) as defined by the *National Clinical Management of HIV Disease Guidelines*[5]. We assessed two primary outcomes:

a. Time to ART initiation based on the difference between the first clinic date and the ART initiation date. If these dates were the same, patients were classified as having same-day ART initiation. ART initiation time/days is reported using six categories (preclinic, same day, 1-7, 8-21, 22-30 and 31+).
b. Time to the first viral load test determined by the duration between the first ART date and the first viral load test date. Time to viral load testing/months is reported using four categories (baseline or pre-ART, 0-6, 7-12 and 13+).

Multivariable logistic regression was used to assess factors associated with ART initiation (31+ days vs. same day initiation) and the uptake of viral load testing (viral load test vs. no viral load test). We report adjusted odds ratios and confidence intervals.

## Results

### Data summary

The analytical sample (Table 1) showed those initiating treatment were more likely to be female (n=4408, 54%), 20-39 years (n=4407, 53%) and located in the South East Region (SERHA (n=4141, 51%). Most were diagnosed at an early HIV stage (n=4430, 60%) and initiated ART at either 31+ days after the first clinic visit (n=3666, 45%) or the same day (n=3463, 43%). The median time to ART initiation was 15 days and most were virally non-suppressed (n=3713, 53%) at the first viral load test. The first viral load tests were mainly done between 0-6 months (n=3047, 37%).

**Table 1.**
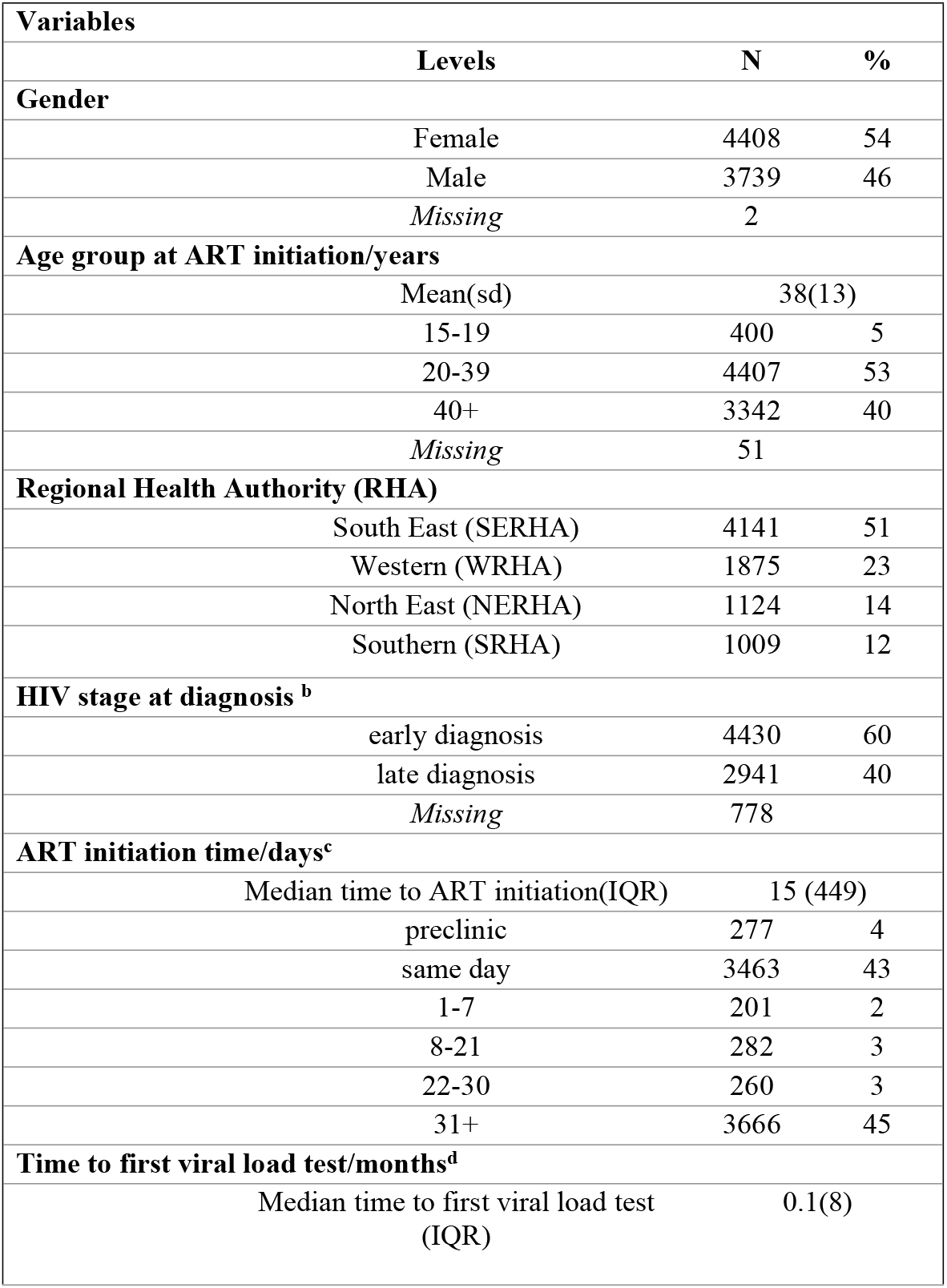
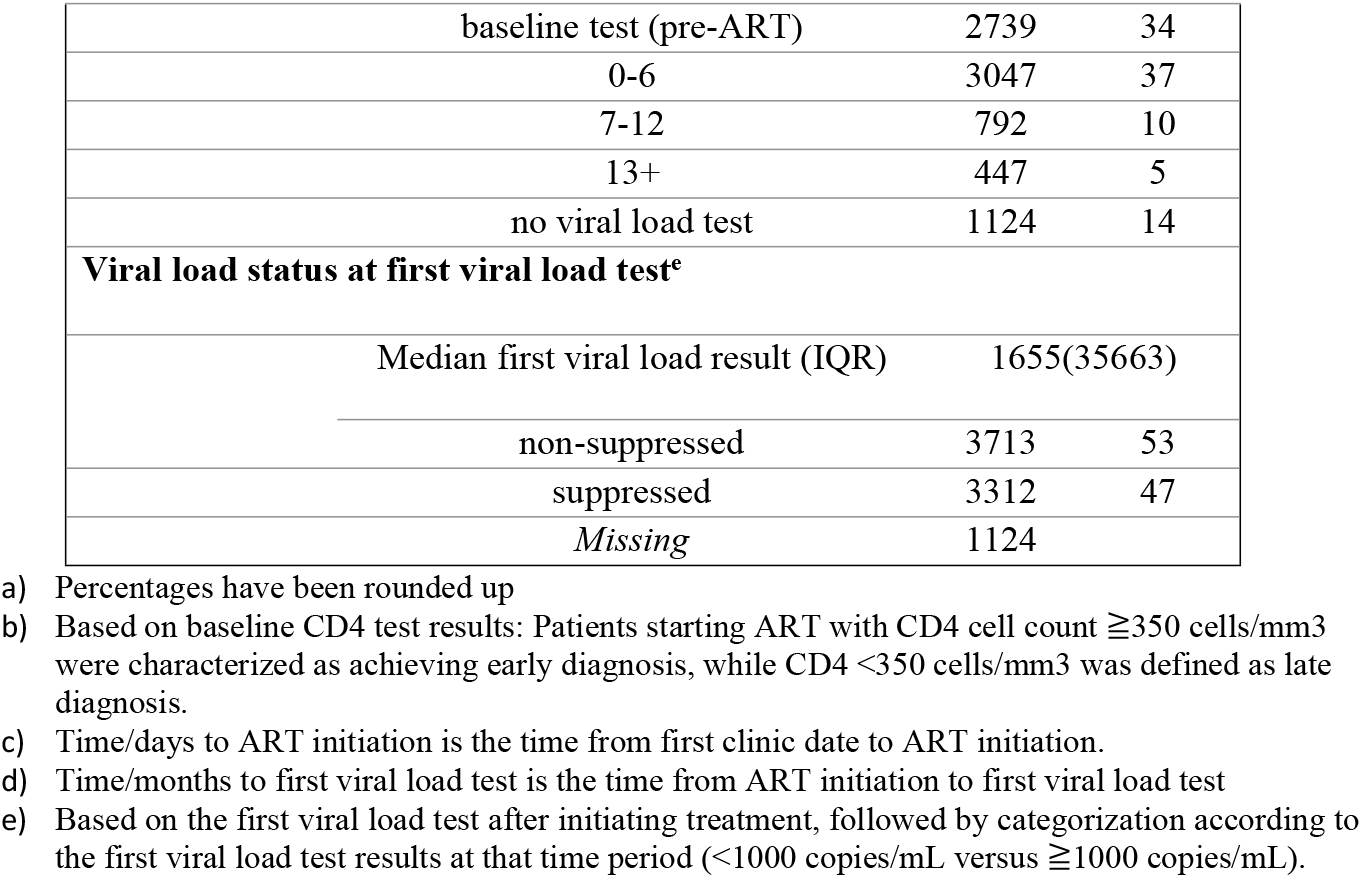
Demographic and clinical characteristics for the analytical sample of PLHIV on HIV treatment, from 2015-2019 in Jamaica (N=8149).

| **Variables** |  |  |  |
| --- | --- | --- | --- |
|  | **Levels** | **N** | **%** |
| **Gender** |  |  |  |
|  | Female | 4408 | 54 |
|  | Male | 3739 | 46 |
|  | *Missing* | 2 |  |
| **Age group at ART initiation/years** |  |  |  |
|  | Mean(sd) | 38(13) |  |
|  | 15-19 | 400 | 5 |
|  | 20-39 | 4407 | 53 |
|  | 40+ | 3342 | 40 |
|  | *Missing* | 51 |  |
| **Regional Health Authority (RHA)** |  |  |  |
|  | South East (SERHA) | 4141 | 51 |
|  | Western (WRHA) | 1875 | 23 |
|  | North East (NERHA) | 1124 | 14 |
|  | Southern (SRHA) | 1009 | 12 |
| **HIV stage at diagnosis <sup>b</sup>** |  |  |  |
|  | early diagnosis | 4430 | 60 |
|  | late diagnosis | 2941 | 40 |
|  | *Missing* | 778 |  |
| **ART initiation time/days<sup>c</sup>** |  |  |  |
|  | Median time to ART initiation(IQR) | 15 (449) |  |
|  | preclinic | 277 | 4 |
|  | same day | 3463 | 43 |
|  | 1-7 | 201 | 2 |
|  | 8-21 | 282 | 3 |
|  | 22-30 | 260 | 3 |
|  | 31+ | 3666 | 45 |
| **Time to first viral load test/months<sup>d</sup>** |  |  |  |
|  | Median time to first viral load test (IQR) | 0.1(8) |  |

|  |  |  |
| --- | --- | --- |
| baseline test (pre-ART) | 2739 | 34 |
| 0-6 | 3047 | 37 |
| 7-12 | 792 | 10 |
| 13+ | 447 | 5 |
| no viral load test | 1124 | 14 |
| **Viral load status at first viral load test<sup>e</sup>** |  |  |
| Median first viral load result (IQR) | 1655(35663) |  |
| non-suppressed | 3713 | 53 |
| suppressed | 3312 | 47 |
| *Missing* | 1124 |  |

- a) Percentages have been rounded up - b) Based on baseline CD4 test results: Patients starting ART with CD4 cell count $\geq 350$ cells/mm<sup>3</sup> were characterized as achieving early diagnosis, while CD4 <350 cells/mm<sup>3</sup> was defined as late diagnosis. - c) Time/days to ART initiation is the time from first clinic date to ART initiation. - d) Time/months to first viral load test is the time from ART initiation to first viral load test - e) Based on the first viral load test after initiating treatment, followed by categorization according to the first viral load test results at that time period (<1000 copies/mL versus $\geq 1000$ copies/mL).

### ART initiation time

Figure 1 shows the proportion of PLHIV with same day ART initiation increased steadily each year from 32% in 2015 to 55% in 2019. This represents a 72% increase over 5 years or a 37.5% increase since the implementation of the “Treat All” strategy in 2017.

**Fig 1.**
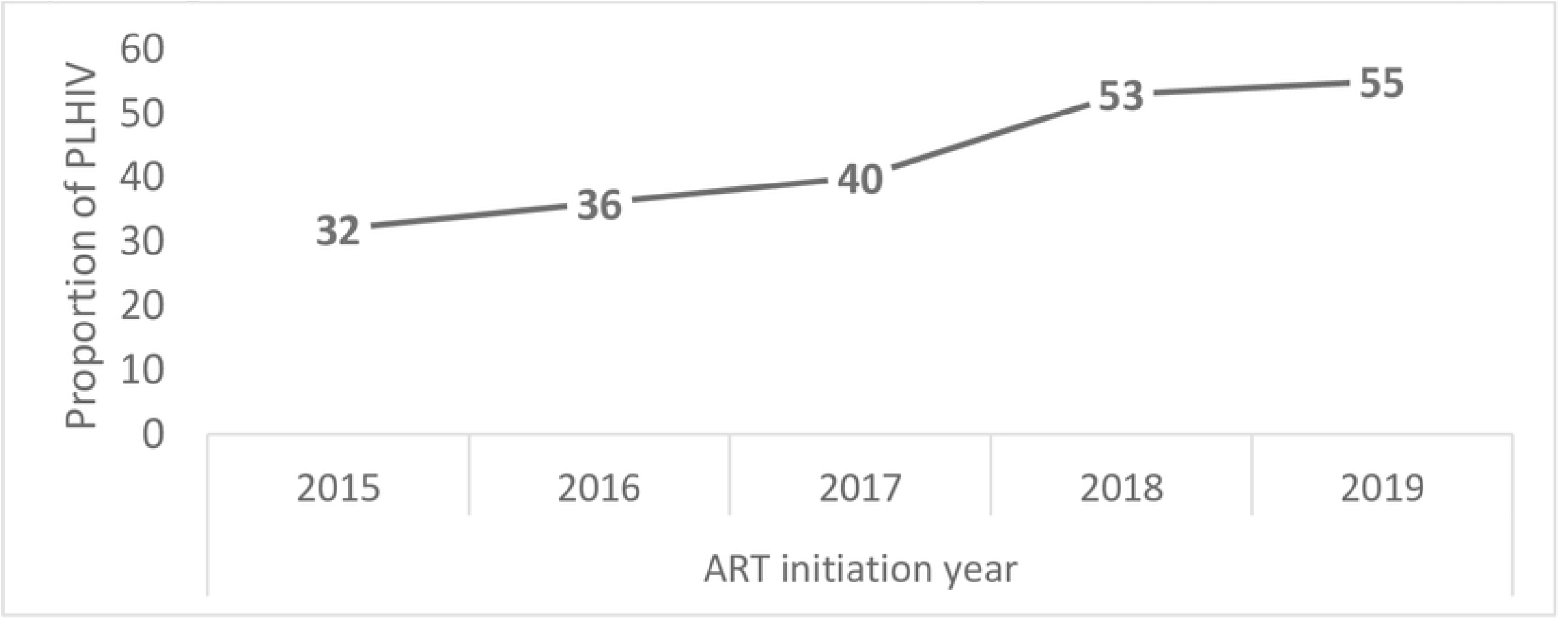
Proportion of PLHIV in Jamaica with same day ART initiation between 2015-2019.

Table 2. shows for persons with same day ART initiation (N= 3461), there was no difference between genders or HIV stage of diagnosis. Most persons were 20-39 years old (n=1817, 53%) and had achieved viral suppression at the first viral load test (n=1477, 53%). For persons initiating ART 31+ days after the first clinic visit (n=3666), most were diagnosed at an early stage (n=2611, 71%) but were virally non-suppressed (n=1965, 58%) at the first viral load test.

**Table 2.**
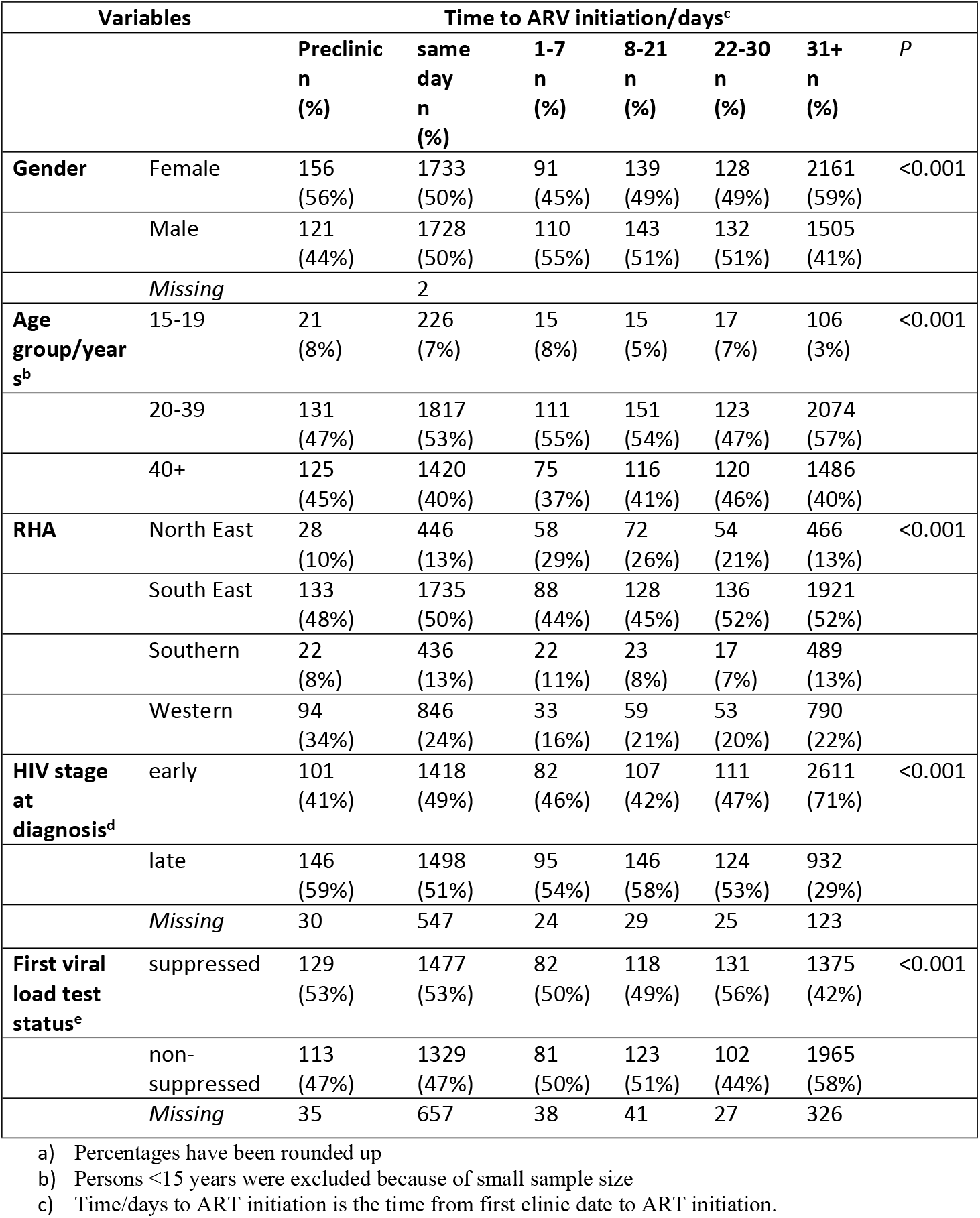
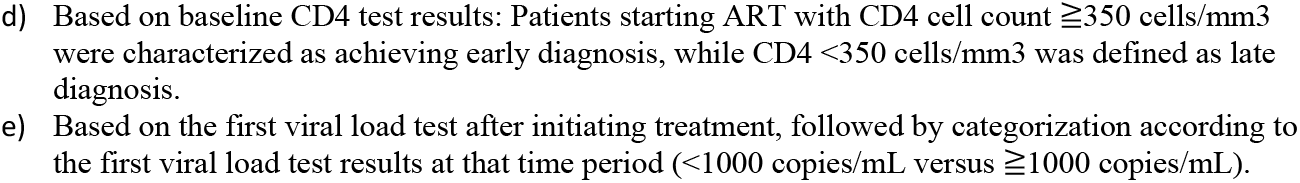
Demographic and clinical characteristics of PLHIV by ART initiation time, for 2015-2019.

### Uptake of viral load testing

Figure 2. shows of persons who were eligible for a 1^st^ viral load test (n=6836), 90% (n=6146) had a test documented and 79% (n=4827) of those had a second viral load test documented. Only 10% (n=690) did not have a first viral load test recorded in TSIS2 despite being eligible to receive testing. Persons were excluded (n=2003) because viral load testing was done before the study period or they were not yet eligible for testing based on the national guidelines.

**Fig 2.**
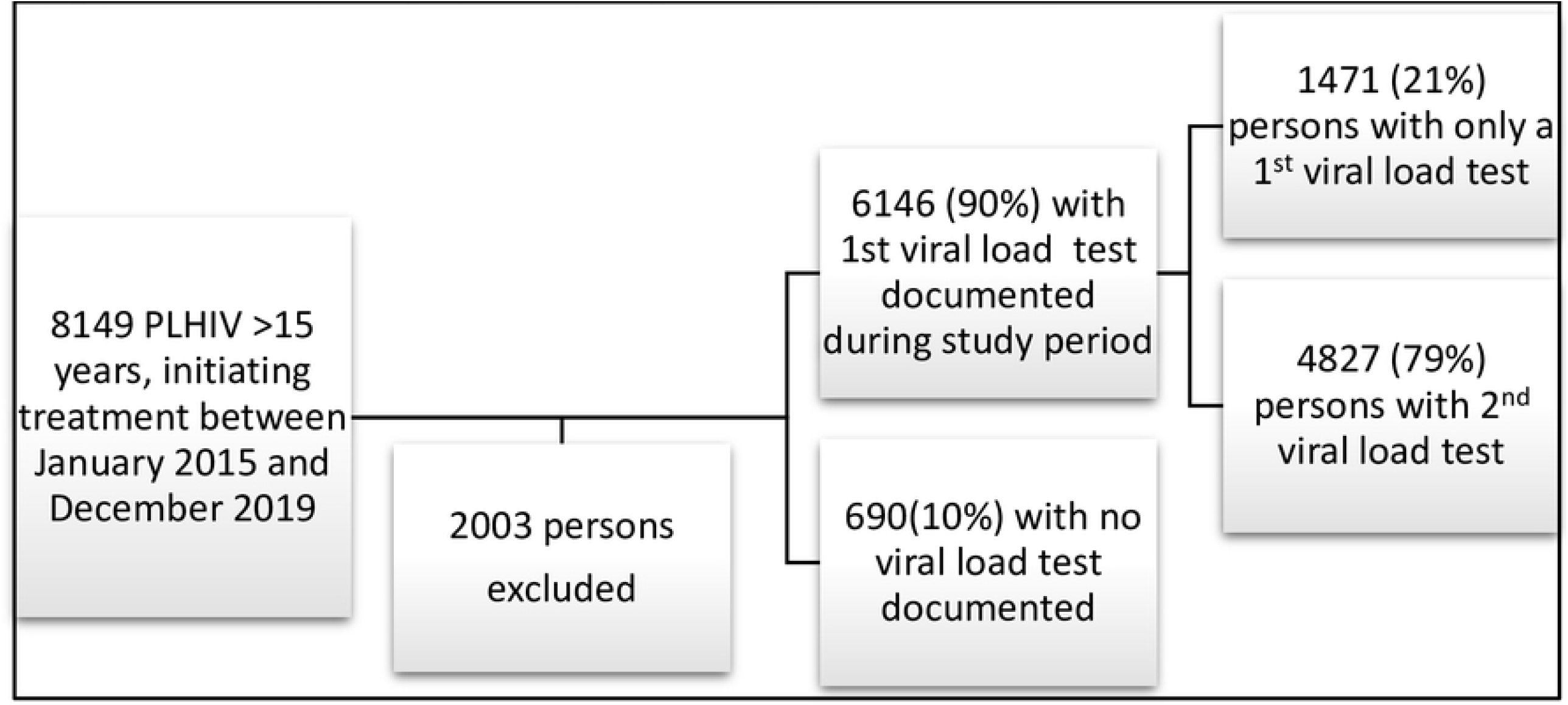
Viral load testing uptake among PLHIV on ART in Jamaica, from 2015-2019.

Table 3. shows among persons with a first viral load test (n=6146), most were female, 20-39 years old at the time of treatment and diagnosed at early HIV stage. Uptake of vl testing was highest in SERHA and lowest in SRHA. Viral load testing was mostly done after 0-6 months on HIV treatment (n=3034, 44%). However, 28% (n=1878) received a baseline vl test before starting treatment. Those with a baseline viral load test were mostly non-suppressed (n=1442, 77%) at the first viral load test. Those without a vl test were mostly males, 20-39 years old, diagnosed early and located in SERHA.

**Table 3.** Demographic and clinical characteristics for PLHIV on HIV treatment by time to uptake of first viral load test.

|  |  | Time to first viral load test/months<sup>b</sup> |  |  |  |  | P |
| --- | --- | --- | --- | --- | --- | --- | --- |
| Variables |  | baseline<br>(pre-ART)<br>n (%) | 0-6<br>n (%) | 7-12<br>n (%) | 13+<br>n (%) | no test<br>n (%) |  |
| **Gender** | Female | 1057<br>(56%) | 1591<br>(52%) | 390<br>(49%) | 217<br>(48%) | 325<br>(47%) | <0.001 |
|  | Male | 821 (44%) | 1443<br>(48%) | 398<br>(51%) | 229<br>(52%) | 364<br>(53%) |  |
|  | Missing |  |  |  |  | 1 |  |
| **Age group/year** | 15-19 | 93 (5%) | 146 (5%) | 38 (5%) | 27 (6%) | 58 (8%) | <0.001 |
|  | 20-39 | 1011(54%) | 1619<br>(53%) | 401<br>(51%) | 256<br>(57%) | 403<br>(58%) |  |
|  | 40+ | 774(41%) | 1269<br>(42%) | 349<br>(44%) | 163<br>(37%) | 229<br>(34%) |  |
| **RHA** | North East | 346 (18%) | 399<br>(13%) | 87<br>(11%) | 45<br>(10%) | 72 (10%) | <0.001 |
|  | South East | 900 (48%) | 1720<br>(57%) | 337<br>(43%) | 210<br>(47%) | 336(48%) |  |
|  | Southern | 268 (14%) | 302<br>(10%) | 75 (9%) | 34 (8%) | 94 (16%) |  |
|  | Western | 364 (20%) | 613<br>(20%) | 289<br>(37%) | 157<br>(35%) | 188<br>(26%) |  |
| **HIV stage at diagnosis<sup>c</sup>** | early | 1105<br>(61%) | 1527<br>(54%) | 411<br>(55%) | 210<br>(49%) | 271<br>(64%) | <0.001 |
|  | late | 711 (39%) | 1326<br>(46%) | 334<br>(45%) | 215<br>(51%) | 150<br>(36%) |  |
| **First viral load test status<sup>d</sup>** | suppressed | 436 (23%) | 1819<br>(60%) | 576<br>(74%) | 226<br>(51%) | NA | <0.001 |
|  | non-suppressed | 1442<br>(77%) | 1215<br>(40%) | 212<br>(26%) | 220<br>(49%) | NA |  |

a) Percentages have been rounded up
b) Time/months to first viral load test is the time from ART initiation to first viral load test
c) Based on baseline CD4 test results: Patients starting ART with CD4 cell count $\geq 350$ cells/mm<sup>3</sup> were characterized as achieving early diagnosis, while CD4 <350 cells/mm<sup>3</sup> was defined as late diagnosis.
d) Based on the first viral load test after initiating treatment, followed by categorization according to the first viral load test results at that time period (<1000 copies/mL versus $\geq 1000$ copies/mL).

### Factors associated with ART initiation and viral load testing uptake

In Figure 3, same day ART initiation was significantly more likely for males compared to females (aOR=1.46, CI=1.32-1.62), 15 - 19 years old compared to 20 - 39 years olds and individuals from WRHA compared to the ones from SERHA. Those virally suppressed were also significantly more likely to be initiated on ART the same day compared to those who were non-suppressed (aOR=1.58, CI=1.43-1.75).

**Fig 3.**
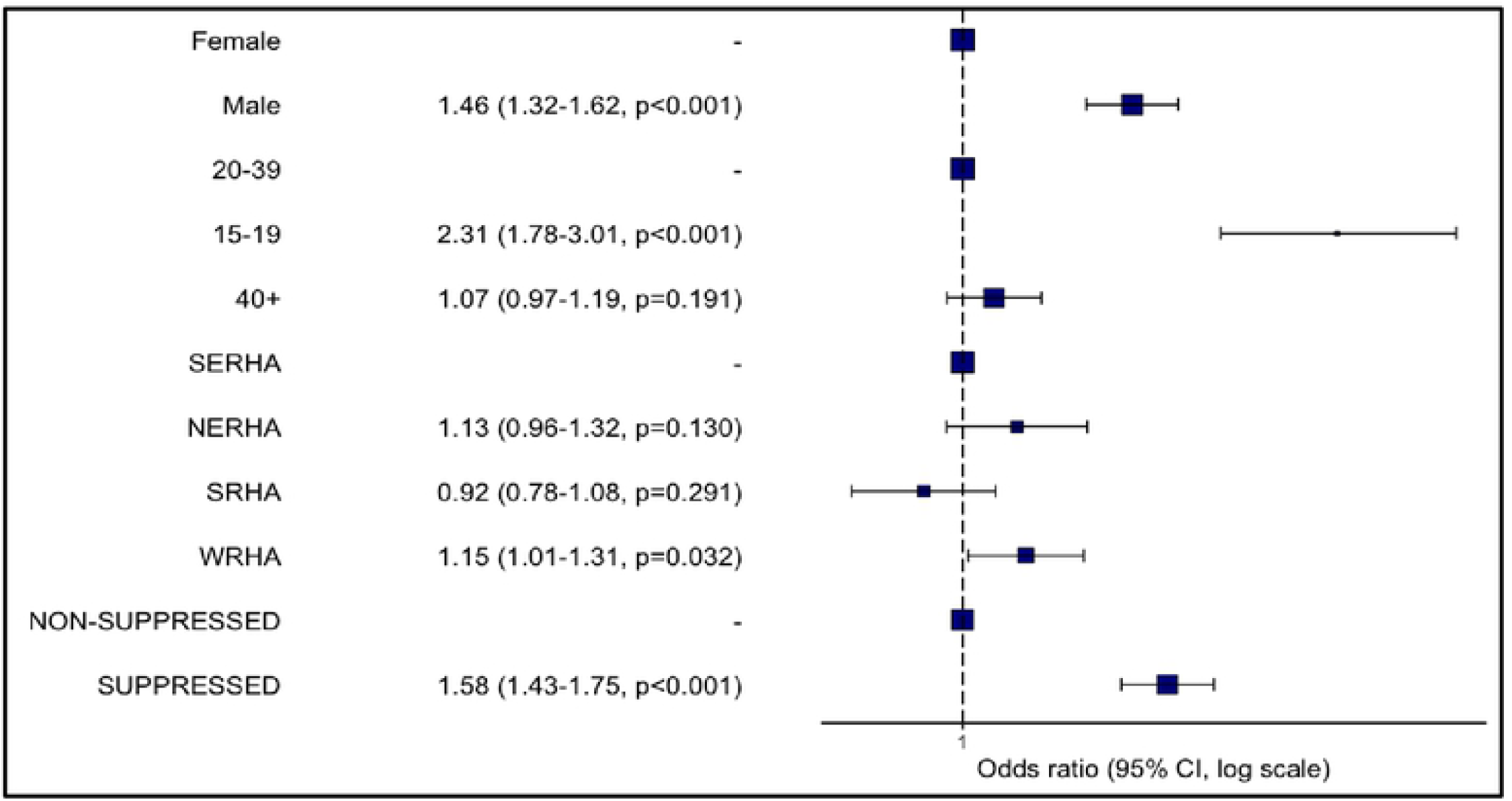
Results of the logistic regression analysis to assess association of PLHIV demographic and clinical variables with ART uptake (31+days vs. same day ART initiation)

Figure 4 shows males were significantly more likely to not have a first viral load test compared to females (aOR=1.55, CI=1.27-1.90). Persons in WRHA were also significantly more likely not to have a viral load test compared to SERHA (aOR=01.29, CI=1.02-1.61). Persons 40+ years were significantly more likely to have a viral load test compared to 20-39 years (aOR=0.73, CI=0.58-0.90). Similarly, persons diagnosed at a late HIV stage (aOR=0.70. CI=0.57-0.86) and those located in NERHA (aOR=0.66, CI=0.41-0.85) where significantly more likely to have a viral load test compared to those diagnosed early and those located in SERHA.

**Fig 4.**
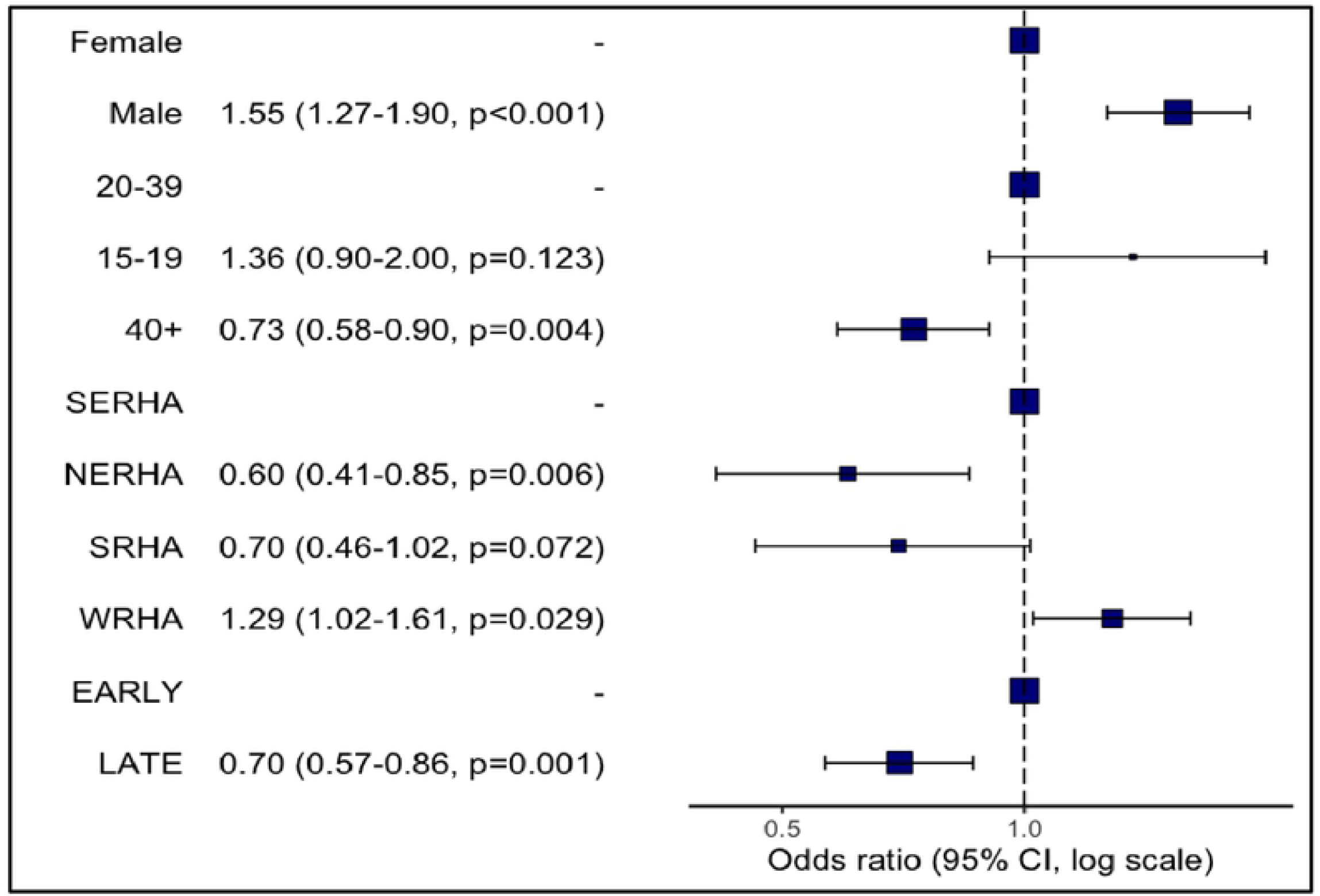
Results of the logistic regression analysis to assess association of PLHIV demographic and clinical variables with viral load status (viral load test vs. no viral load test)

## Discussion

Our study shows the current goal of immediate ART initiation is gradually being met, demonstrated by a 72% increase in same day ART initiation over the five-year study period. Those initiating treatment were mostly females but males were significantly more likely to have same day initiation. Persons initiating ART later (31+ days after the first clinic visit) were mostly virally non-suppressed while those with same day initiation had achieved viral suppression at the first viral load test. This demonstrates the benefit of immediate HIV treatment. However, while immediate treatment initiation is the current goal, it is important to ensure persons are also ready to start treatment [5]. Evidence suggests rapid initiation of ART could lead to increased loss to follow-up because of insufficient time to accept and disclose HIV status and to prepare for lifelong treatment[7].

Coverage of the first viral load test was 90% among eligible persons but declined to 79% for the second test, indicating Jamaica has adapted routine viral load testing but retention is lacking. For viral suppression to occur once on treatment, continuous engagement with care is necessary[14]. WHO estimates on average, between 64% to 94% of people will still be in care one year after starting HIV treatment[15]. Considering the frequency of missing viral load tests, there is need to assess the barriers to retention and conduct routine assessment of patients with missing records to provide more robust information for quality improvement and to mitigate possible service delivery challenges.

Most persons were virally non-suppressed at first viral load test and had received this test between 0-6 months after initiating ART, which is consistent with the current national guidelines[5]. Coverage of baseline (pre-ART initiation) viral load testing was 28%, despite the national guidelines stipulating testing after persons have started treatment. Males and persons with late HIV diagnosis were significantly more likely to not have a viral load test. Males have demonstrated the tendency to delay in seeking health care[16] and persons with a late diagnosis may also be more likely to delay viral load testing. Jamaica has a decentralized health system but HIV viral load testing is only provided by the national laboratory, which limits access. UNAIDS estimates that globally 95% of HIV service delivery is currently facility based and to optimize efficiencies, it is suggested that community-based service delivery be ramped up[17]. In a previous paper[18], we suggest Jamaica explore scaling up viral load testing through decentralizing testing to treatment sites, some of which are community based. Research has also demonstrated how differentiated service delivery (DSD) can be applied to improve uptake in specific populations, such as males[19,20]. Successful DSD implementation aims to simplify and adapt services to patients needs and requires that countries determine the barriers to access as well as specific needs of hard-to-reach populations to improve uptake of services[21] and consequently, health outcomes.

We also recommend exploring the effects of supply chain performance on ART and viral load testing uptake. A better understanding of these consumption variables at service delivery points can only serve to improve patient management and bridge the analysis gap. This has become particularly relevant within the context of the COVID-19 pandemic which saw HIV services being diverted or slowed resulting in stock outs and backlogs in viral load testing[22]. It should not be assumed that poor uptake only occurs as a result of a patient’s negligence when there may be challenges within the system or supply chain[23].

We recognize the limitations of the data. Improved accuracy is expected if the data was normally distributed[24,25]. A recent data quality audit carried out by the HIV Strategic Information Unit measured the completeness of data in TSIS2 at 63%[12]. Missing data or non-entry of data could be responsible for persons not having a documented viral load test and not necessarily non-uptake of testing services. The data did not allow us to measure the time retained on ART, time retained in care or assess virologic failure. We are unable to relate timing of viral load test to clinic attendance or adherence. Table 3. shows 23% of persons were virally suppressed prior to ART initiation, indicating these persons may have been in treatment at clinics not included in the government data (or overseas). The data includes patients receiving both private and public care and did not attempt to separate. However, the sample size is large and includes all persons on HIV treatment for the study period, which limits selection bias. According to the c-statistic from the regression analyses, the variables assessed produced strong models with 62% accuracy for viral load testing uptake and 60% accuracy for ART initiation. Medians are reported to account for skewness of the data.

## Conclusion

The goal of immediate ART initiation is increasingly being met and is significantly associated with viral suppression at the first viral load test. Males were more likely to be initiated on ART the same day as the first clinic visit but less likely to have a viral load test after ART initiation. A qualitative assessment should be conducted to understand important challenges in accessing routine viral load testing, followed by implementation of differentiated care models targeting males.

## Data Availability

Data can be obtained from the Ministry of Health and Wellness, Jamaica-National HIV Strategic Information Unit.

## Financial Disclosure statement

The author(s) received no specific funding for this work.

## Acknowledgements

The authors would like to thank the HIV/STI/TB Strategic Information Unit for providing the baseline data.

## Acronyms

ART – antiretroviral therapy,

PLHIV – persons living with HIV,

NERHA – Northeast Regional Health Authority,

RHA- Regional Health Authority,

SERHA-Southeast Regional Health Authority,

SRHA – Southern Regional Health Authority,

WRHA – Western Regional Health Authority.

## Data Sharing Statement

The study investigator was required to sign a confidentiality agreement which prevents data sharing. Full data can be requested directly from the Ministry of Health and Wellness.

## Notes

### Competing Interest Statement

The authors have declared no competing interest.

### Funding Statement

No funding was received for this research

### Author Declarations

The Ethics Committee of the Ministry of Health of Jamaica gave ethical approval for this work under study number 20/2017.

